# Analysis and Forecast of COVID-19 Pandemic in Pakistan

**DOI:** 10.1101/2020.06.24.20138800

**Authors:** Abdul Bari Malik

## Abstract

The COVID-19 infections in Pakistan are spreading at an exponential rate and a point may soon be reached where rigorous prevention measures would need to be adopted. Mathematical models can help define the scale of an epidemic and the rate at which an infection can spread in a community. I used ARIMA Model, Diffusion Model, SIRD Model and Prophet Model to forecast the magnitude of the COVID-19 pandemic in Pakistan and compared the numbers with the reported cases on the national database. Results depicts that Pakistan could hit peak number of infectious cases between June 2020 and July, 2020.

## 1 Introduction

The first case of COVID-19 emerged in Pakistan on 26th February 2020 in Karachi, the most populous city of Pakistan. The patient zero had a travel history to Iran and immediately quarantined upon testing positive for the virus. However, the patient was followed by hundreds of pilgrims returning from Iran which were likely carrying the virus that ultimately led to the spread of COVID-19 into the community. Since then the infections have been increasing exponentially and without proper intervention the situation may escalate enough to overwhelm the already struggling healthcare system in the country. In this study we aim to model the COVID-19 pandemic in Pakistan, which will indicate the peak infection day, rate of increase of infections per day and a 90-day forecast to assess the final size of the epidemic.

## 2 Models Used

The accuracy of traditional forecasting largely depends on the availability of data to base its predictions and estimates of uncertainty. In outbreaks of pandemics there is no data at all in the beginning and then limited as time passes, making predictions widely uncertain.

To forecast confirmed cases of COVID-19, we adopt time series forecasting approaches. I am using models from the ARIMA family, SIR family, PROPHET Models and Diffusion Gradient family.

### 2.1 ARIMA (Autoregressive integrated moving average)

ARIMA, short for **‘Autoregressive Integrated Moving Average’** is actually a class of models that ‘explains’ a given time series based on its own past values, that is, its own lags and the lagged forecast errors, so that equation can be used to forecast future values. Any ‘non-seasonal’ time series that exhibits patterns and is not a random white noise can be modeled with ARIMA models. Since COVID-19 data for Pakistan depicts some **‘non-stationarity’** and ‘**non-seasonal’** pattern in it and reflects a linearity among data points. That’s why ARIMA is our first choice.

#### ARIMA model in words

▪ Predicted Yt = Constant + Linear combination Lags of Y (upto p lags) + Linear Combination of Lagged forecast errors (upto q lags)

If in future our time series will reflect some seasonal pattern, then we would be need to add seasonal terms and it becomes SARIMA, short for ‘Seasonal ARIMA’

### 2.2 Diffusion Process

I am using a model from a paper by Emmanuelle Le Nagard and Alexandre Steyer, that attempts to reflect the social structure of a diffusion process. The model is also sensitive to when we define the origin of time for the epidemic process. Here, I just took the first point of the time series available. I am exploring the 3d parameter space to find a minimum, using Gradient Descent.

#### Basic Model for Diffusion

▪ Y (t) = α(m − Y (t))
  ○ where (α > 0) plus the “typical” initial condition Y (0) = 0.
  ○ Y denotes the cumulative number of adopters at time t
  ○ m the size of the population of potential adopters
  ○ α the intensity of the spreads

### 2.3 SIRD Model of Epidemiology for COVID-19

Since the emergence of COVID-19, several mathematical models have been developed to simulate the rate of infection spread, infections per day and the resolution of the epidemic. The SIRD model refers to the number of susceptible, infected, resolved and deceased cases during an epidemic at any given time. The model assumes that susceptible cases (S), infected cases (I), resolved cases (R) and Deceased cases (D) are compartments and each individual of a given population will pass through the susceptible phase then to the infected phase and finally to the recovered phase.

The SIRD model is a steady state model, therefore the population that is analyzed is static i.e. no one is being born or is dying. Additionally, the model assumes that once a person is infected, they are immune to the disease and therefore cannot contract it again. The SIRD model is ideal for modelling the spread of diseases spread through person to person contact.

#### Basic SIRD Model

▪ Number of people who are (stocks):
  ○ S_t_ = Susceptible
  ○ I_t_ = Infectious
  ○ R_t_ = Resolving
  ○ D_t_ = Dead
  ○ C_t_ = ReCovered
▪ Constant population size is N
  ○ S_t_ + I_t_ + R_t_ + D_t_ + C_t_ = N
▪ Susceptible get infected at rate β_t_I_t_/N
  ○ New infections = β_t_I_t_/N · S_t_
▪ Infectiousness resolve at Poisson rate γ, so the average number of days that a person is infectious is 1/γ so γ = .2 ⇒ 5 days
▪ Post-infectious cases then resolve at Poisson rate θ. E.g. θ = .1 ⇒ 10 days
▪ Resolution happens in one of two ways:
  ○ Death: fraction δ
  ○ Recovery: fraction 1 – δ
▪ SIRD Model - Laws of Motion
  ○ ΔSt+1 = −β_t_S_t_I_t_ / N (new infections)
  ○ ΔIt+1 = β_t_S_t_I_t_/N (new infections) − γI_t_ (resolving infectious)
  ○ ΔRt+1 = γI_t_ (resolving infectious) − θR_t_ (cases that resolve)
  ○ ΔDt+1 = δθR_t_ (die)
  ○ ΔCt+1 = (1 − δ)θR_t_ (recovered)
  ○ D0 = 0

### 2.4 Prophet

Prophet is a procedure for forecasting time series data based on an additive model where non-linear trends are fit with yearly, weekly, and daily seasonality, plus holiday effects. It works best with time series that have strong seasonal effects and several seasons of historical data. Prophet is robust to missing data and shifts in the trend, and typically handles outliers well. Prophet automatically evaluates forecast performance and flags issues that warrant manual intervention.

#### Prophet uses two approaches for saturating forecasts

▪ Forecasting Growth
▪ Saturating Minimum

Prophet uses a linear model for its forecast. When forecasting growth, there is usually some maximum achievable point: total market size, total population size, etc. This is called the carrying capacity, and the forecast should saturate at this point. Prophet also allows you to make forecasts using a logistic growth trend model, with a specified carrying capacity.

For COVID-19 Pakistan forecast, we are using Linear Model. But I have space of implementing a logistic growth function and lag parameter for obtaining Logistic growth with Prophet.

#### Basic Prophet Model

▪ Prophet is an additive model with the following components:
  ○ y(t) = g(t) + s(t) + h(t) + ϵ_t_
  ○ g(t) models trend, which describes long-term increase or decrease in the data. Prophet incorporates two trend models, a saturating growth model and a piecewise linear model, depending on the type of forecasting problem.
  ○ s(t) models seasonality with Fourier series, which describes how data is affected by seasonal factors such as the time of the year (e.g. more searches for eggnog during the winter holidays)
  ○ h(t) models the effects of holidays or large events that impact business time series (e.g. new product launch, Black Friday, Superbowl, etc.)
  ○ ϵ_t_ represents an irreducible error term

## 3 Data

The data was extracted from daily situation reports published on the National Institute of Health (NIH) Pakistan for a period of 120 days, (from February 26, 2020 to June 20, 2020) and was corroborated with the simulation results

## 4 Analysis

The NIH Pakistan data for the cumulative cases and daily reported cases was plotted to observe the increase in number of cases for a period of 116 days.

Figure (4.1) shows total count of Confirmed Cases, Fatalities and Recovered Cases at provincial level since the pandemic started in Pakistan. The current trajectory suggests an exponential increase in the number of cases since the pandemic started in Pakistan on 26th February 2020 (Figure 4.2). The data on the number of daily reported cases also reflects a general increasing trend as testing for COVID-19 gathers pace (Figure 4.3).

**Figure 4-1:**
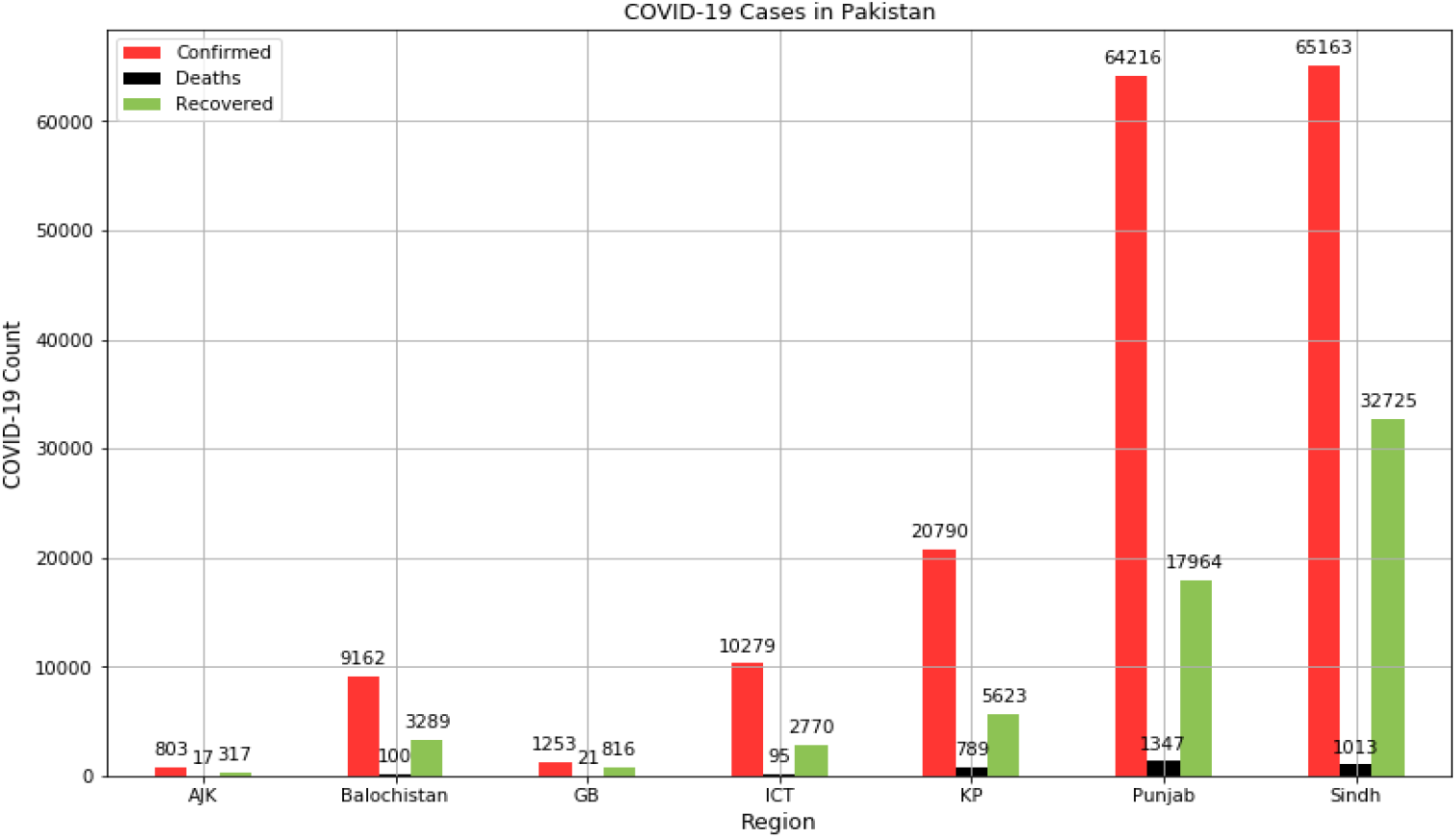
The COVID-19 cases in Pakistan since the outbreak began on 26th February 2020.

**Figure 4-2:**
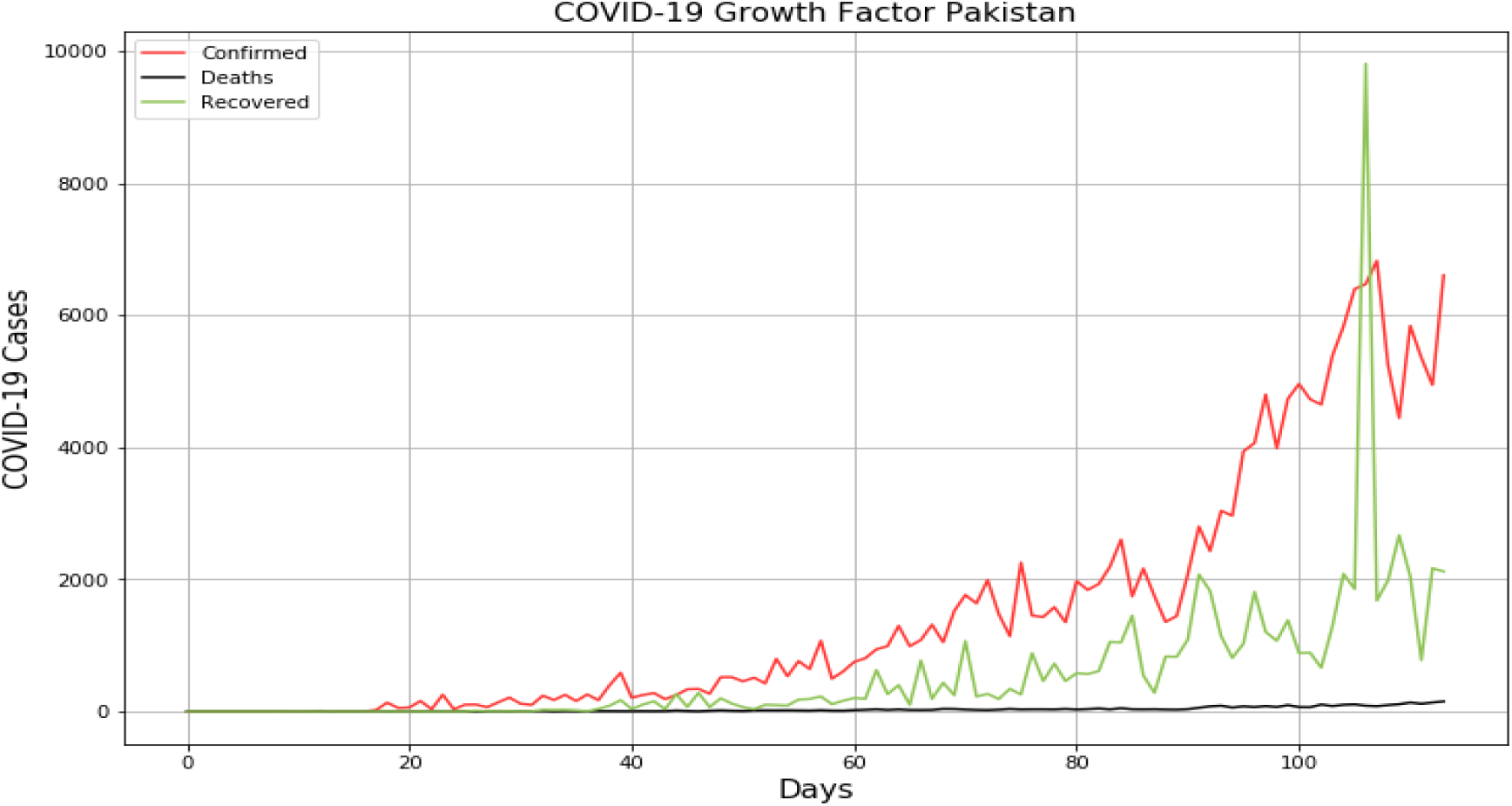
The graph shows an exponential growth pattern for the 116-day data extracted from the NIH, Pakistan database. The x-axis corresponds to the days, whereas the y-axis corresponds to the number of cases.

**Figure 4-3:**
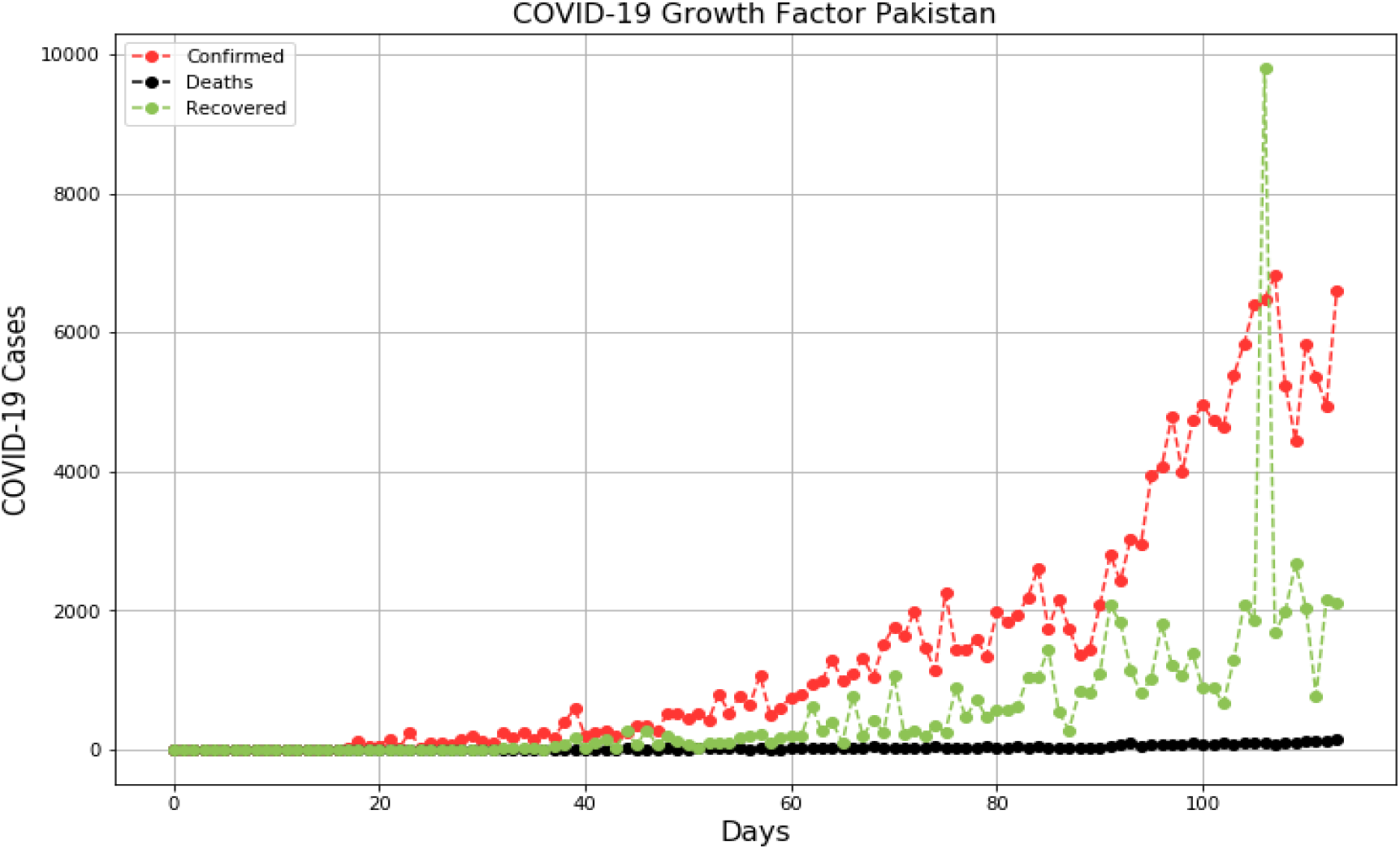
Daily reported COVID-19 cases in Pakistan since the outbreak on 26th February 2020. The graph shows an exponential growth pattern for the - day data extracted from the NIH, Pakistan database. On day 1 of the epidemic the number of cases were 2 and on day 116 the number of daily reported cases were 6604. These numbers may represent the gap in the unreported or asymptomatic cases in Pakistan. The x-axis corresponds to the days, whereas the y-axis corresponds to the number of cases.

**Figure 4-4:**
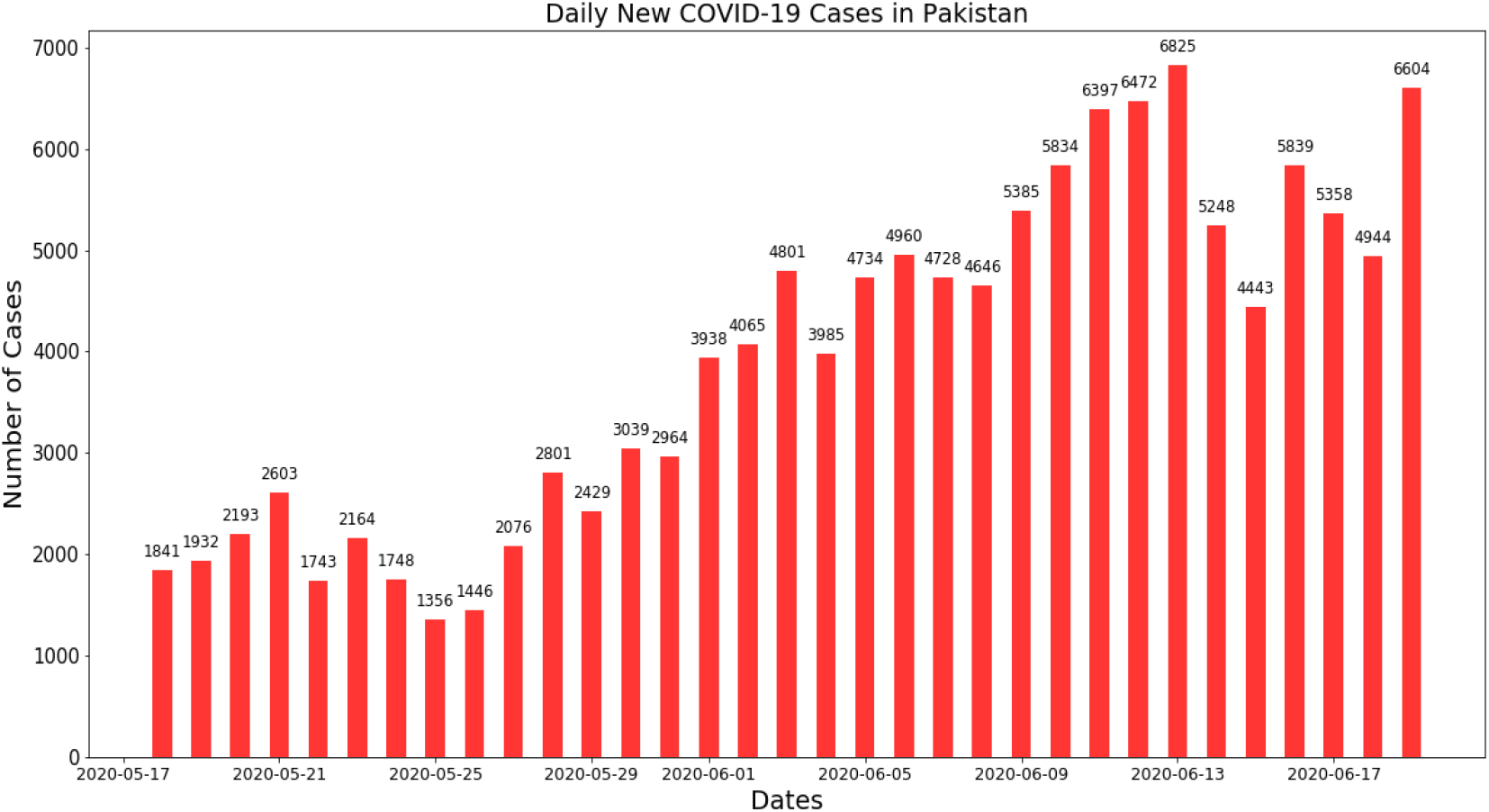
Daily Reported Cumulative Tests Positive (Daily Confirmed Cases) in Pakistan.

Figures (4.4), (4.5) and (4.6) depicts daily count of last 30 days for confirmed cases, fatalities and recovered cases respectively. While Figures (4.7) and (4.8) reflects weekly increase in number of confirmed cases and fatalities in Pakistan. Figure (4.9) depicts monthly increase in number of confirmed cases. Fatalities and recovered cases in Pakistan.

**Figure 4-5:**
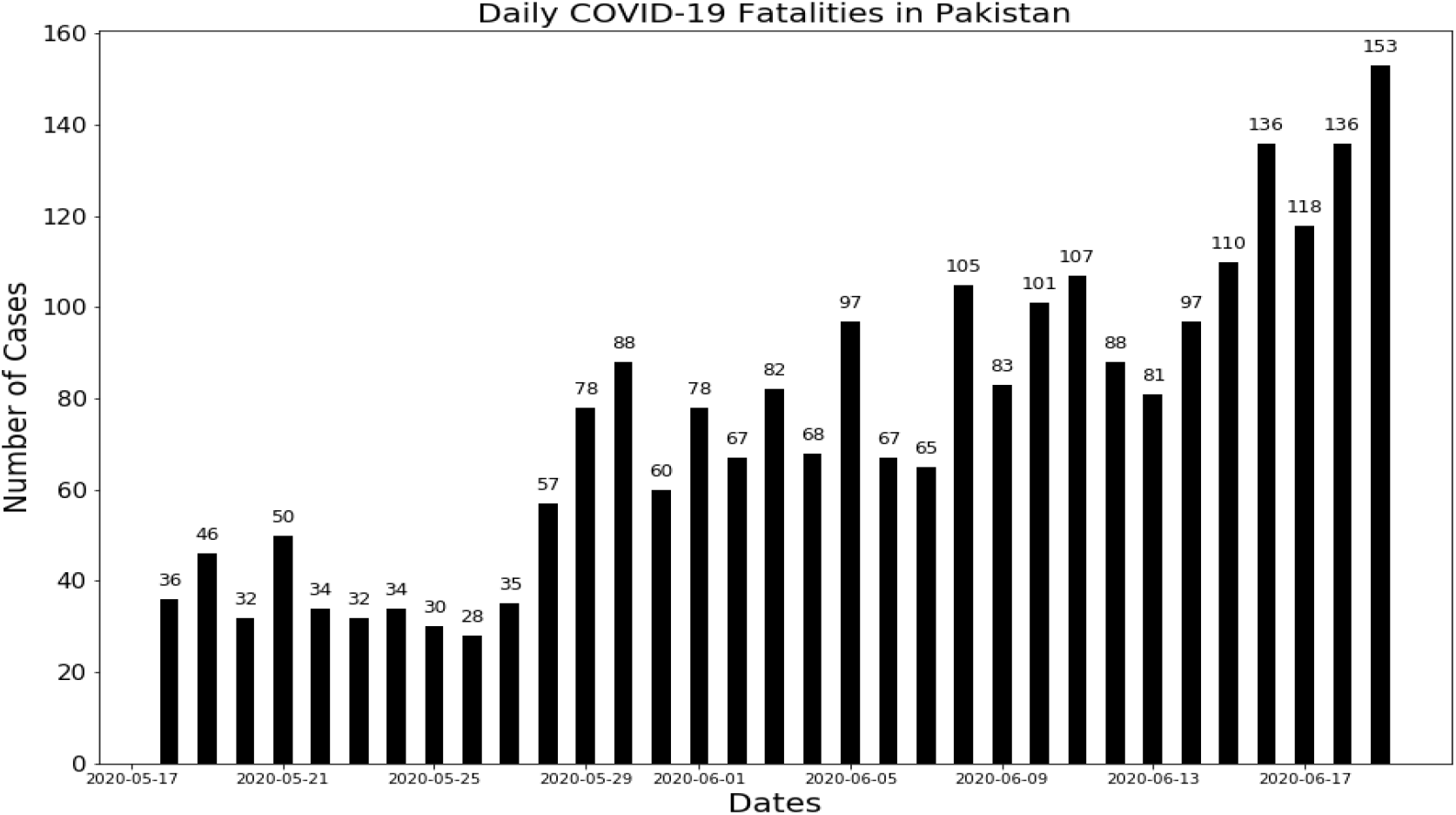
Daily reported Cumulative COVID-19 Fatalities in Pakistan.

**Figure 4-6:**
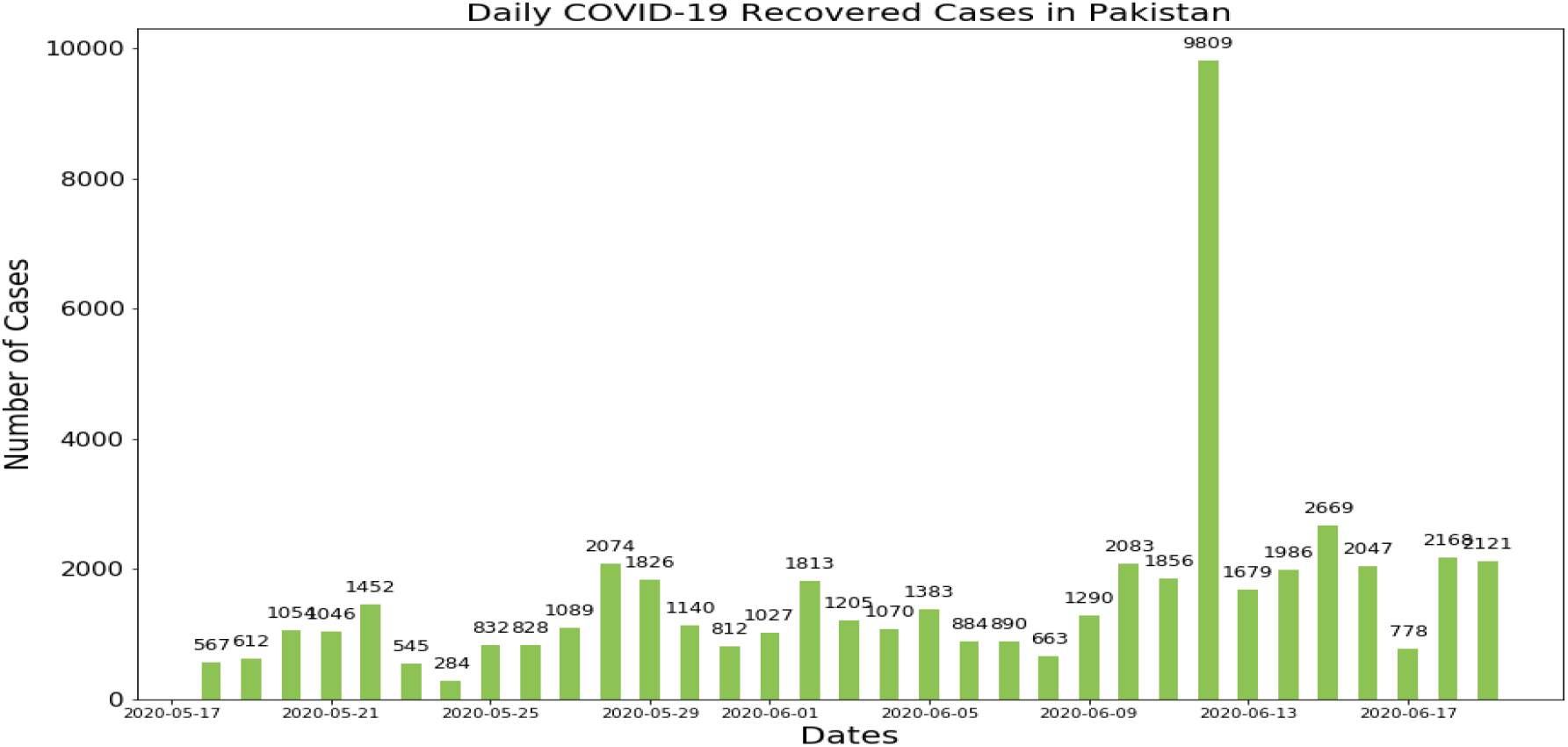
Daily Reported Recovered Cases in Pakistan.

**Figure 4-7:**
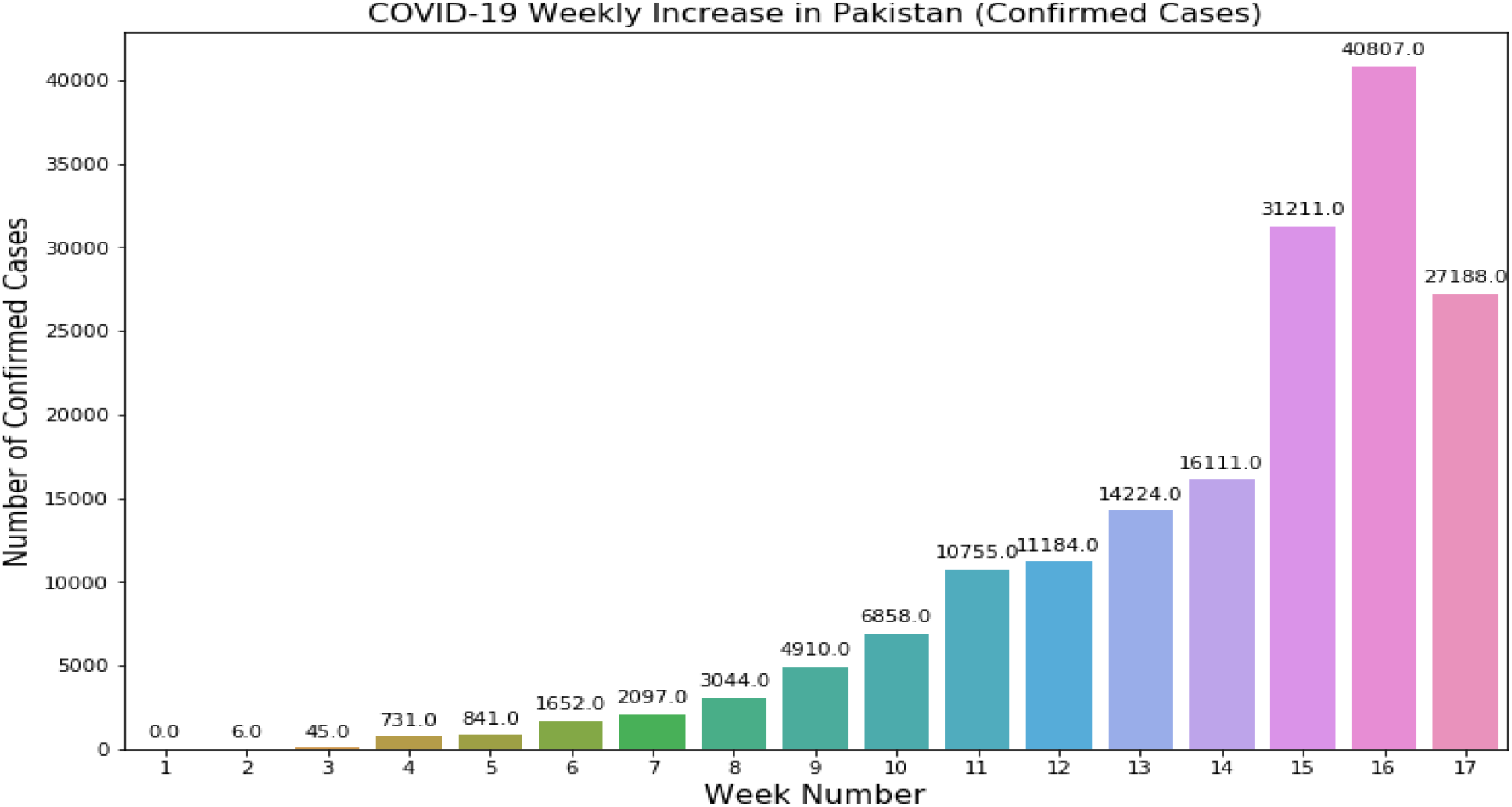
Weekly increase in number of Confirmed Cases in Pakistan. The figure shows reported Cumulative Tests Positive (Daily Confirmed Cases) per week since the outbreak began on 26th February 2020.

**Figure 4-8:**
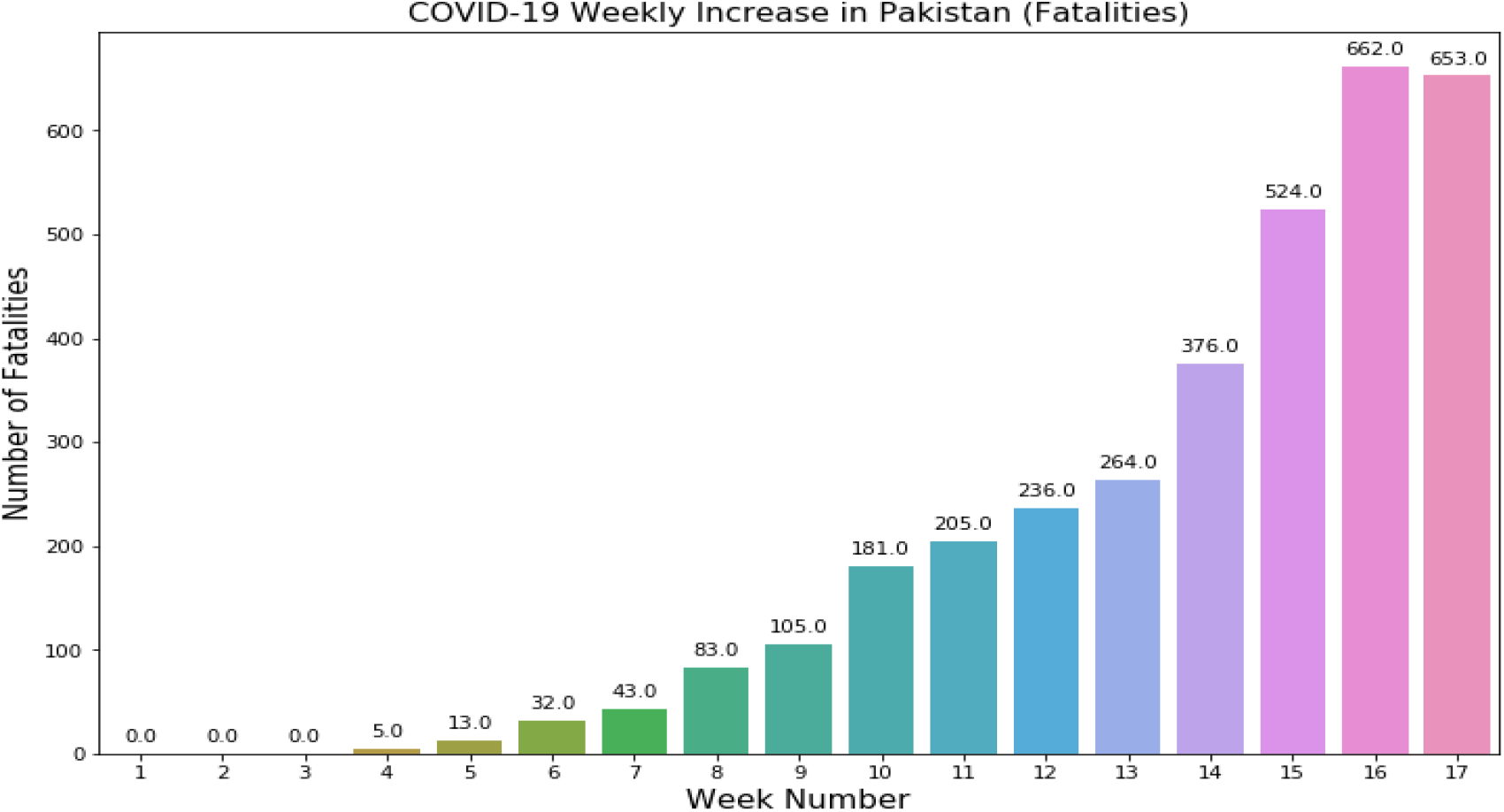
Weekly increase in number of Fatalities in Pakistan. The Figure shows increase in number of fatalities per week since the outbreak began on 26th February 2020.

**Figure 4-9:**
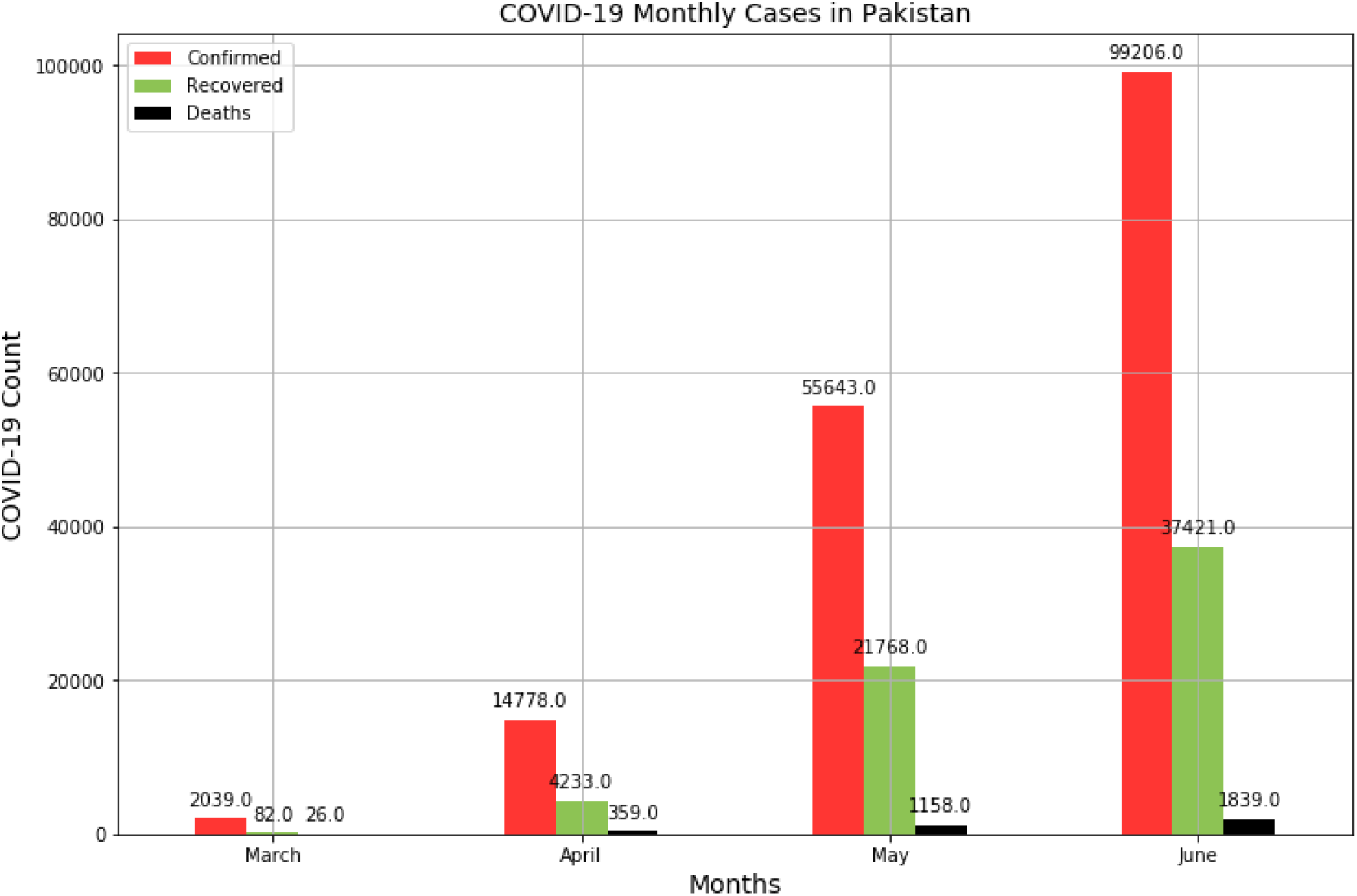
Cumulative Monthly COVID-19 cases in Pakistan. The Figure depicts monthly increase in number of positive test cases, fatalities and recovered personnel in Pakistan.

## 5 Projections for PAKISTAN

**Figure 5-1:**
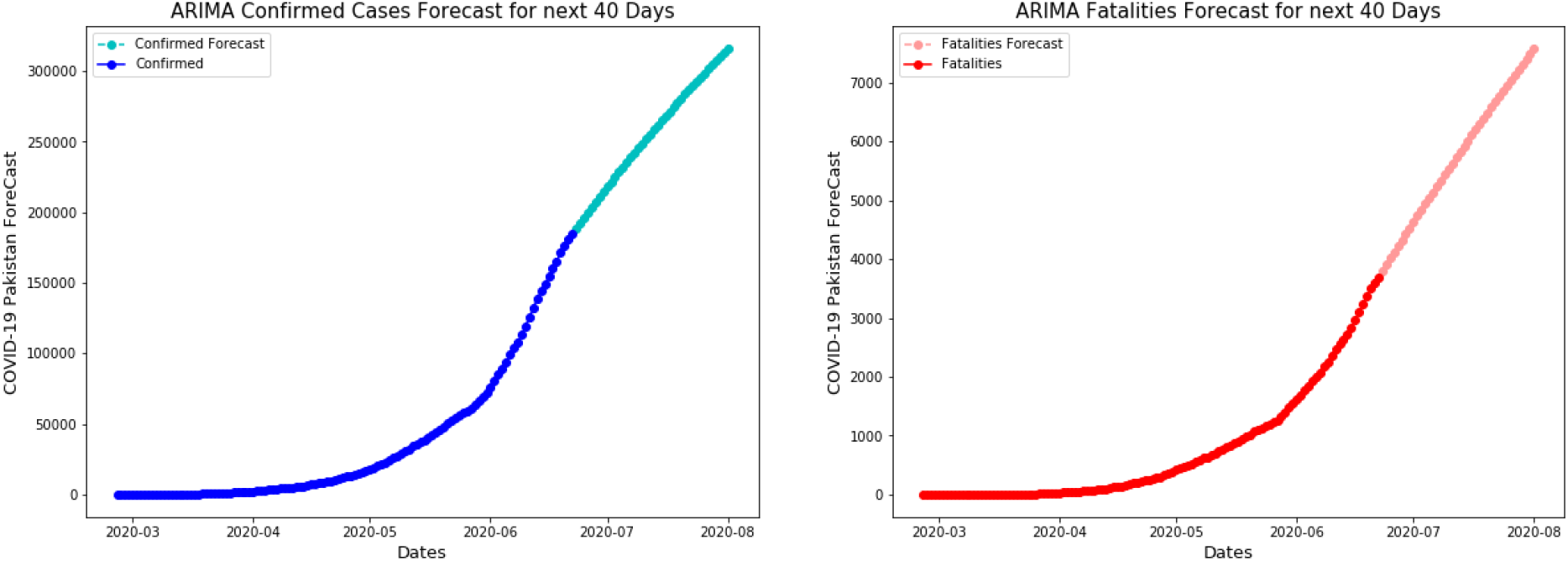
COVID-19 estimation using ARIMA. COVID-19 forecast depicts that COVID-19 Confirmed cases numbers will rise to the count of 300,000 and deceased count will rise to the number 7000 by August 2020.

**Figure 5-2:**
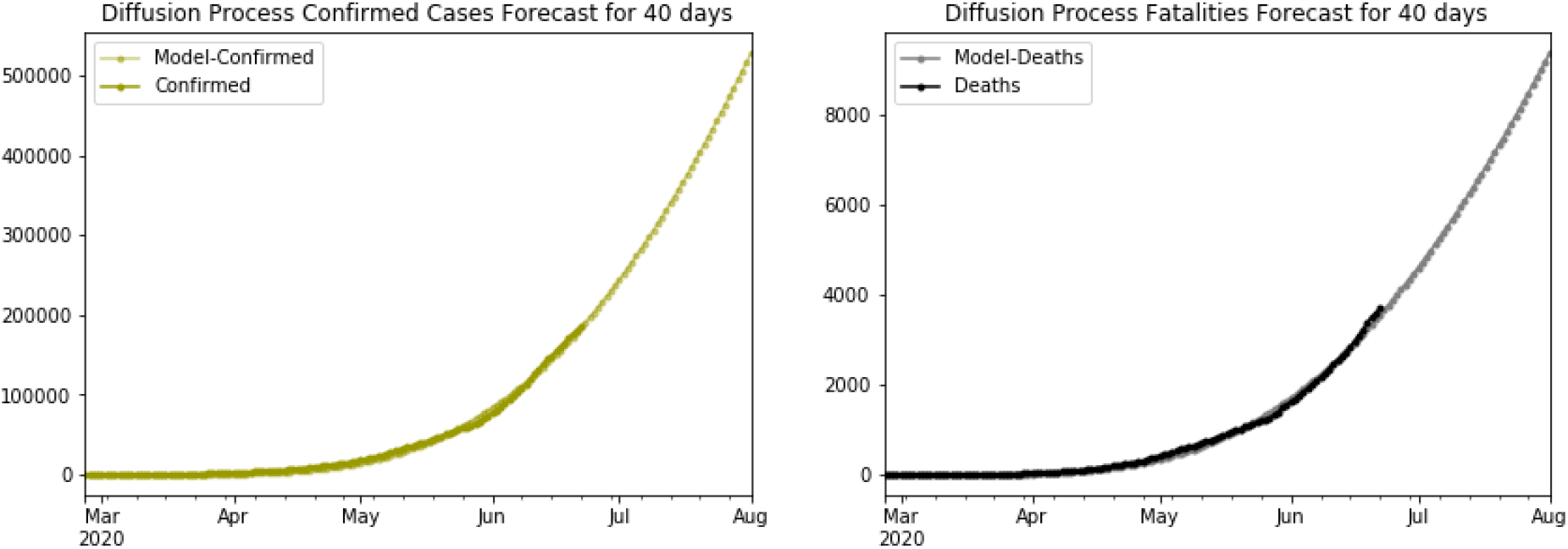
COVID-19 Projections using Diffusion Process. COVID-19 forecast illustrates that COVID-19 Confirmed cases numbers will rise to the count of 500,000 and deceased count will rise above number 8000 by August 2020.

**Figure 5-3:**
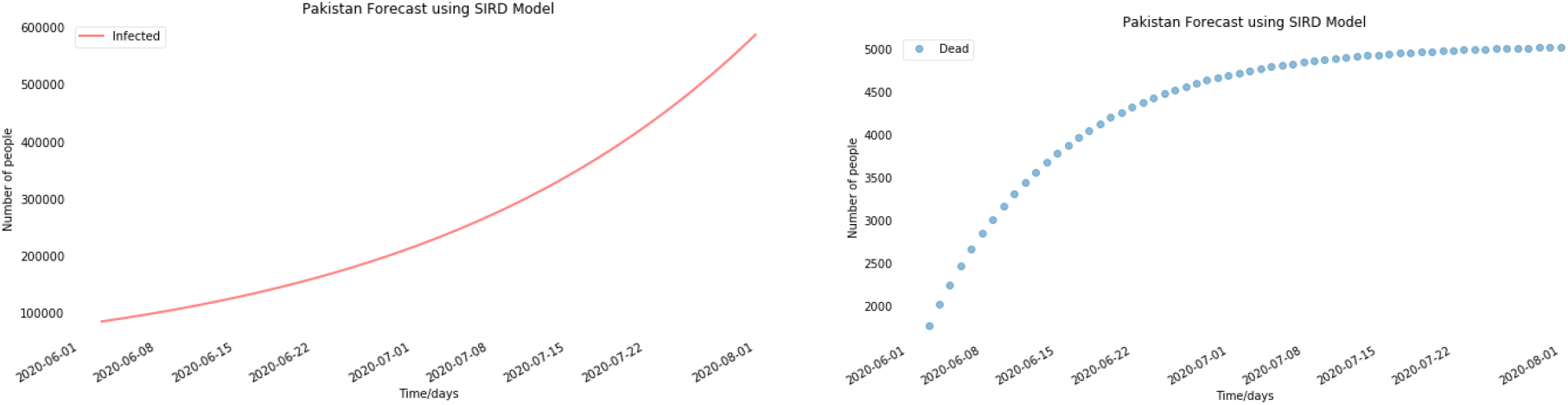
COVID-19 Projections using SIRD. COVID-19 forecast shows that COVID-19 Confirmed cases numbers will rise to the count of 600,000 and deceased tally will rise above figures of 5000 by August 2020.

**Figure 5-4:**
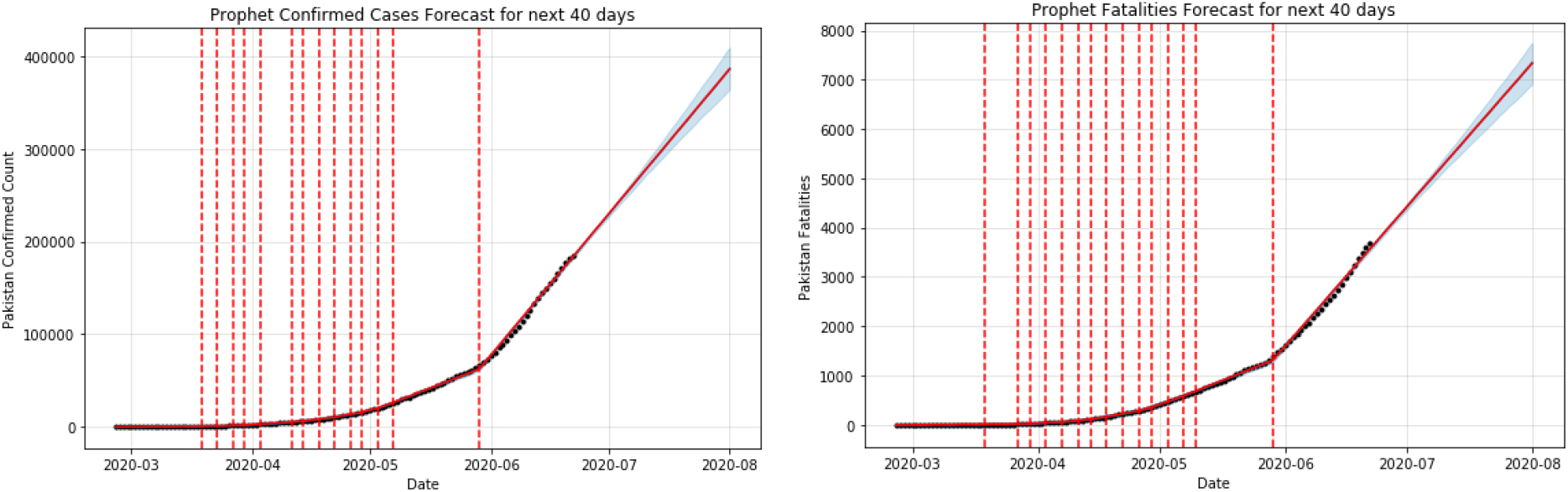
COVID-19 Projections using Prophet. COVID-19 forecast depicts that COVID-19 Confirmed cases numbers will rise to the tally of 600,000 and deceased count will rise above figures of 5000 by August 2020.

**Figure 5-5:**
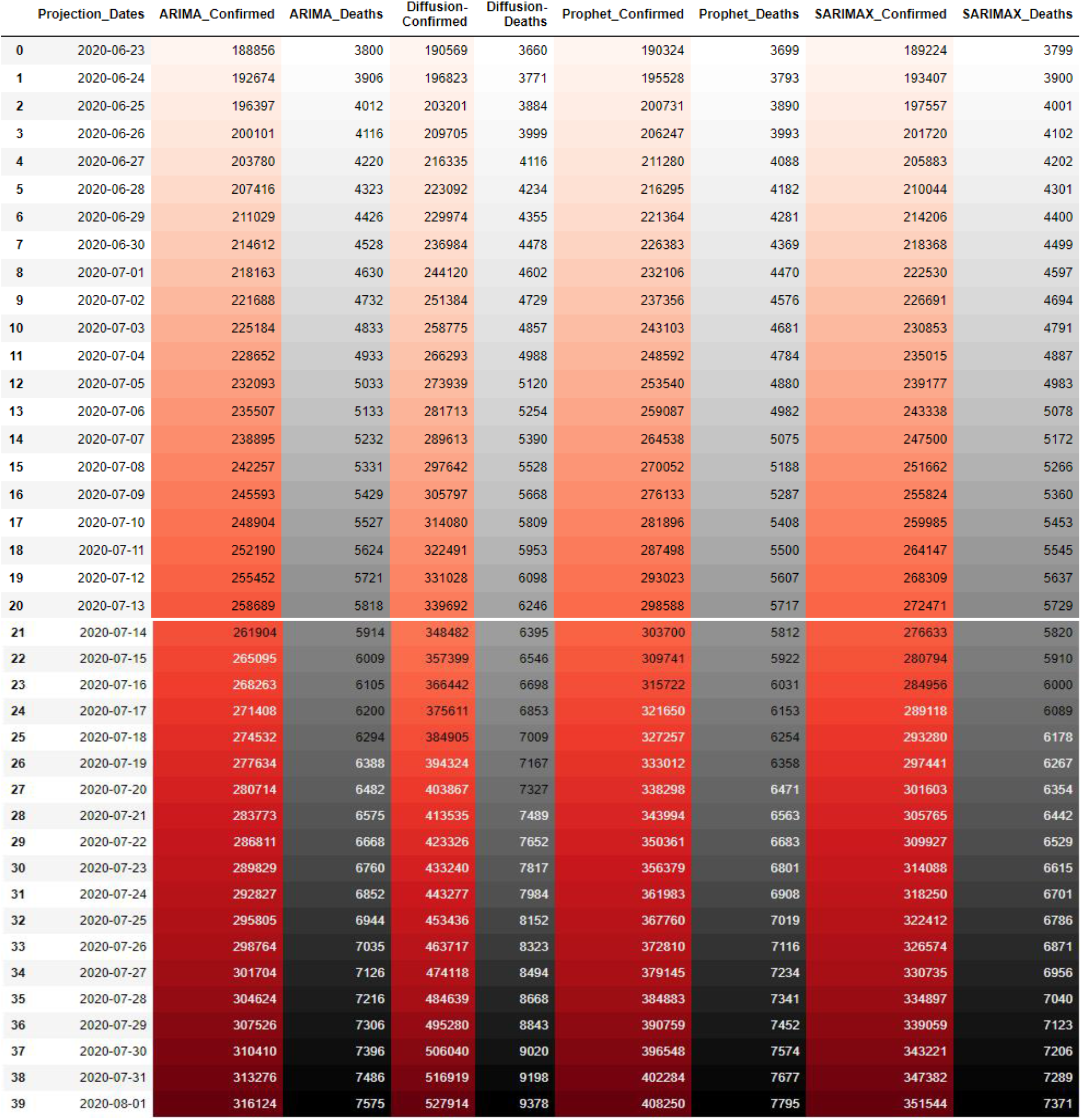
COVID-19 Projections for Pakistan: Table shows 40 days projections using ARIMA family, Diffusion and Prophet.

## 6 Conclusion

As the epidemiological data suggests, a huge number of population is susceptible to be affected by COVID-19 infection. The government has already closed the educational institutions and the partial lockdown has been imposed in the country. Efforts should be made to reduce the spread of the virus through social distancing, contact tracing, isolation and quarantine measures. Most importantly, massive testing should be done in the potential population to point out the silent spreader of the virus to reduce the disease spread. There should be a strong communication along with data and resource sharing between central and provincial authorities. Our frontline army is now our healthcare professionals. Steps should be taken to reduce the risk of healthcare providers being infected from the patient by providing recommended personal protective equipment (PPE).

## Data Availability

Data for COVID-19 pandemic in Pakistan available on government of Pakistan dashboard for COVID-19.

http://covid.gov.pk/stats/pakistan

